# Digital Divide or Educational Divide? The Impact of Social Media on COVID-19 Vaccination in Middle-Aged and Older Adults

**DOI:** 10.1101/2025.09.05.25335178

**Authors:** Mason Dupre, Truls Østbye, Teah M. Bayless, Hanzhang Xu

## Abstract

The spread of disinformation and harmful content on social media is well-known. However, most research focuses on the negative influence of social media on the well-being of adolescents and teenagers. We used data from the 2022 Health and Retirement Study (n=4,038) to examine how the use of social media (i.e., Facebook, Twitter [X], and Instagram) was associated with vaccination uptake for Covid-19 among U.S. middle-aged and older adults. Frequency of social media use was measured on a scale ranging from 0=never to 5=daily. In our study (mean (SD) age: 69.4 (9.7) years), approximately 35.9% of participants used social media daily while 34.3% never did. Multivariable logistic regression models showed that social media use was not directly associated with Covid-19 vaccination. However, there was a significant interaction between social media use and educational attainment. Among adults who frequently used social media, those with low education were significantly less likely to be vaccinated for Covid-19 compared to their more educated counterparts (*P* for interaction =.008). These results remained after accounting for differences in participants’ sociodemographic background, health status, religiosity, and other factors. Furthermore, social media use was not associated with vaccination for influenza, pneumonia, or shingles. Our results highlight the importance of delivering tailored public health messages when promoting vaccine uptake among less-educated older populations.

## INTRODUCTION

The widespread use of social media platforms such as Facebook, X (formerly Twitter), and Instagram has increasingly transformed how individuals access and consume health information.^1,2^ Although numerous studies have documented the impact of social media on the health and well-being of adolescents and young adults,^3–6^ much less is known about how social media use influences adults at older ages. The Covid-19 pandemic was an especially notable period that amplified the consumption of social media in growing segments of the adult population seeking to maintain social connections and obtain health-related information.^1,7^ However, these platforms also exposed users to inaccurate/deceptive content and misleading public health information.

Prior studies have shown that individuals with lower levels of educational attainment are more susceptible to health misinformation across a wide range of health conditions.^8–10^ Studies have also shown that individuals with inadequate (or inaccurate) health literacy are less likely to get vaccinations or follow other health behavior recommendations.^11,12^ The current research report used data from a nationally-representative sample of U.S. adults aged 50 and older to examine whether the frequency of social media use influenced the uptake of Covid-19 vaccination and whether educational attainment had an impact on the likelihood of vaccination.

## METHODS

### Data

The analysis used nationally-representative data of U.S. middle-aged and older adults from the Health and Retirement Study (HRS). The HRS is sponsored by the National Institute on Aging (grant number: U01AG009740) and is conducted by the University of Michigan.^13^ The HRS is the largest ongoing longitudinal study of adults over age 50 that has interviewed more than 40,000 participants since it was launched in 1992.^13,14^ The current study used data from 4,284 respondents aged 50 and older who participated in the 2022 Leave-Behind Questionnaire that assessed the use of social media. Respondents with missing data on study variables were omitted (n=246, 5%). The final analytical sample included 4,038 HRS participants.

### Measures

#### Vaccinated for Covid-19

Participants were asked if they ever had a Covid-19 vaccine (yes or no). Two participants reported not knowing if they were vaccinated and were coded as “no.” Overall, 89.5% of HRS participants reported that they had been vaccinated and 10.5% reported that they had not been vaccinated (standard deviation [SD]=0.31). Participants were also asked if they had been vaccinated for influenza (75.1% yes; SD=0.43), pneumonia (43.4% yes; SD=0.50), and/or shingles (29.1% yes; SD=0.45).

#### Social Media Use

To assess social media use, participants were asked how often they accessed social network sites like Facebook, Twitter [X], or Instagram. Responses ranged on a scale from 0 (never) to 5 (daily). Overall, 34.3% of HRS respondents reported never using social media and 35.9% reported daily use (SD=1.7).

#### Covariates

We included measures for age (years), sex (male or female), race and ethnicity (non-Hispanic White, non-Hispanic Black, Hispanic, or non-Hispanic other race), region of residence (Northeast, Midwest, South, or West), educational attainment (years), marital status (never married, married, divorced, or widowed), religious attendance (1+ times per year, 2-3 times per month, once a week, or more than weekly), frequency of computer use (continuous scale 1-7), frequency of using the internet for health information (never, once a year, once a month, several times a week, or daily), and reported doctor-diagnoses of high blood pressure, diabetes mellitus, heart disease, and cancer (yes or no; for each condition).

### Statistical Analysis

We examined the distributions of participants’ sociodemographic, behavioral, and health-related characteristics for the overall sample and by Covid-19 vaccination status using chi-square and Kruskal-Wallis tests for categorical and continuous variables, respectively. Multivariable logistic regression models were then used to examine the association between frequency of social media use and Covid-19 vaccination status. We tested an interaction term between social media use and educational attainment to examine whether the association between social media use and vaccination was modified by years of education. Finally, we assessed whether the associations were observed for reported vaccination for influenza, pneumonia, and/or shingles. All analyses were performed using Stata 18.0 (StataCorp LP, College Station, TX). P values < 0.05 were considered statistically significant.

## RESULTS

The mean age of study participants was 69.4 years (SD=9.7), 59.3% were female, and the majority were non-Hispanic White adults (61.3%) (Table 1). Overall vaccination rates for Covid-19 were high (89.5%; SD=0.31) and participants who were older, married, and had a history of chronic disease were more liked to be vaccinated. Table 2 presents the results from the logistic regression models showing the association between frequency of social media use and Covid-19 vaccination status. Social media use was not directly associated with being vaccinated for Covid-19 in the unadjusted (Model 1) or fully-adjusted model (Model 2). However, a significant interaction emerged between social media use and educational attainment (Model 3). Among adults who frequently used social media, those with low education were significantly less likely to be vaccinated for COVID-19 compared to their more educated counterparts (*P* for interaction =.008). Figure 2 illustrates the differences in the participants’ predicted probability of Covid-19 vaccination by frequency of social media use and years of education (estimated from Model 3). Additional analyses (Supplementary Tables 1-3) further showed that social media use was not significantly associated with inoculation for other reported vaccines, including influenza, pneumonia, and shingles.

**Table 1.** Characteristics of Study Participants by Covid-19 Vaccination, Health and Retirement Study, 2022 (n=4,038)

|  | Total<br>(n=4,038) | Never<br>Vaccinated<br>(n=424) | Ever<br>Vaccinated<br>(n=3,614) | *P*<br>value |
| --- | --- | --- | --- | --- |
| Age, mean (SD), y | 69.4 (9.7) | 67.0 (10.1) | 69.7 (9.6) | <0.001 |
| Sex |  |  |  | 0.480 |
| Female | 2,393 (59.3%) | 258 (60.8%) | 2,135 (59.1%) |  |
| Male | 1,645 (40.7%) | 166 (39.2%) | 1,479 (40.9%) |  |
| Race and Ethnicity |  |  |  | 0.018 |
| NH White | 2,477 (61.3%) | 279 (65.8%) | 2,198 (60.8%) |  |
| NH Black | 785 (19.4%) | 69 (16.3%) | 716 (19.8%) |  |
| Hispanic | 580 (14.4%) | 48 (11.3%) | 532 (14.7%) |  |
| NH Other | 196 (4.9%) | 28 (6.6%) | 168 (4.6%) |  |
| Region of Residence |  |  |  | 0.001 |
| Northeast | 595 (14.7%) | 35 (8.3%) | 560 (15.5%) |  |
| Midwest | 809 (20.0%) | 91 (21.5%) | 718 (19.9%) |  |
| South | 1,663 (41.2%) | 184 (43.4%) | 1,479 (40.9%) |  |
| West | 971 (24.0%) | 114 (26.9%) | 857 (23.7%) |  |
| Education, mean (SD), y | 13.5 (3.0) | 13.1 (2.6) | 13.5 (3.0) | 0.015 |
| Marital Status |  |  |  | 0.030 |
| Never married | 320 (7.9%) | 37 (8.7%) | 283 (7.8%) |  |
| Married | 2,279 (56.4%) | 211 (49.8%) | 2,068 (57.2%) |  |
| Divorced | 753 (18.6%) | 95 (22.4%) | 658 (18.2%) |  |
| Widowed | 686 (17.0%) | 81 (19.1%) | 605 (16.7%) |  |
| Religious Attendance |  |  |  | <0.001 |
| Never | 1,611 (39.9%) | 168 (39.6%) | 1,443 (39.9%) |  |
| 1+ times per year | 696 (17.2%) | 55 (13.0%) | 641 (17.7%) |  |
| 2-3 times per month | 403 (10.0%) | 26 (6.1%) | 377 (10.4%) |  |
| Once a week | 887 (22.0%) | 107 (25.2%) | 780 (21.6%) |  |
| More than weekly | 441 (10.9%) | 68 (16.0%) | 373 (10.3%) |  |
| Frequency of Computer Use (1-7) | 3.4 (2.4) | 3.2 (2.5) | 3.4 (2.4) | 0.037 |
| Use Internet for Health Info. |  |  |  | 0.090 |
| Never | 557 (13.8%) | 70 (16.5%) | 487 (13.5%) |  |
| Once a year | 741 (18.4%) | 73 (17.2%) | 668 (18.5%) |  |
| Once a month | 1,521 (37.7%) | 171 (40.3%) | 1,350 (37.4%) |  |
| Several times a week | 866 (21.4%) | 84 (19.8%) | 782 (21.6%) |  |
| Daily | 353 (8.7%) | 26 (6.1%) | 327 (9.0%) |  |
| Social Media Use |  |  |  | 0.430 |
| Never | 1,384 (34.3%) | 144 (34.0%) | 1,240 (34.3%) |  |
| Once a year | 161 (4.0%) | 13 (3.1%) | 148 (4.1%) |  |
| Once a month | 338 (8.4%) | 28 (6.6%) | 310 (8.6%) |  |
| Several times a week | 705 (17.5%) | 75 (17.7%) | 630 (17.4%) |  |
| Daily | 1,450 (35.9%) | 164 (38.7%) | 1,286 (35.6%) |  |
| Diagnosed Conditions: |  |  |  |  |
| High blood pressure | 2,512 (62.2%) | 233 (55.0%) | 2,279 (63.1%) | 0.001 |
| Diabetes mellitus | 1,158 (28.7%) | 101 (23.8%) | 1,057 (29.2%) | 0.019 |
| Heart disease | 1,035 (25.6%) | 103 (24.3%) | 932 (25.8%) | 0.500 |
| Cancer | 685 (17.0%) | 51 (12.0%) | 634 (17.5%) | 0.004 |

Abbreviations: SD, standard deviation; NH, non-Hispanic. Note: Values reported as frequency (percentage) or mean (SD).

**Table 2.** Model Estimates of the Association Between Social Media Use, Education, and Vaccination for Covid-19, HRS 2022 (n=4,038)

|  | **Model 1** |  | **Model 2** |  | **Model 3** |  |
| --- | --- | --- | --- | --- | --- | --- |
|  | OR [95% CI] | *P*<br>value | OR [95% CI] | *P*<br>value | OR [95% CI] | *P*<br>value |
| Frequency of Social Media Use | 0.97 [0.92-1.03] | .358 | 0.98 [0.92-1.05] | .560 | 0.70 [0.53-0.91] | .007 |
| Years of Education |  |  | 1.05 [1.01-1.09] | .013 | 1.00 [0.94-1.06] | .881 |
| Interaction Term |  |  |  |  |  |  |
| Social Media Use * Education |  |  |  |  | 1.03 [1.01-1.05] | .008 |

Abbreviations: HRS, Health and Retirement Study; OR, odds ratio; CI, confidence interval.
*Note:* Models 2 and 3 adjusted for age, sex, race, ethnicity, geographic region, marital status, religious attendance, frequency of computer use, frequency of internet use for health information, and reported diagnoses of high blood pressure, diabetes, heart disease, and cancer.

**Figure 1.**
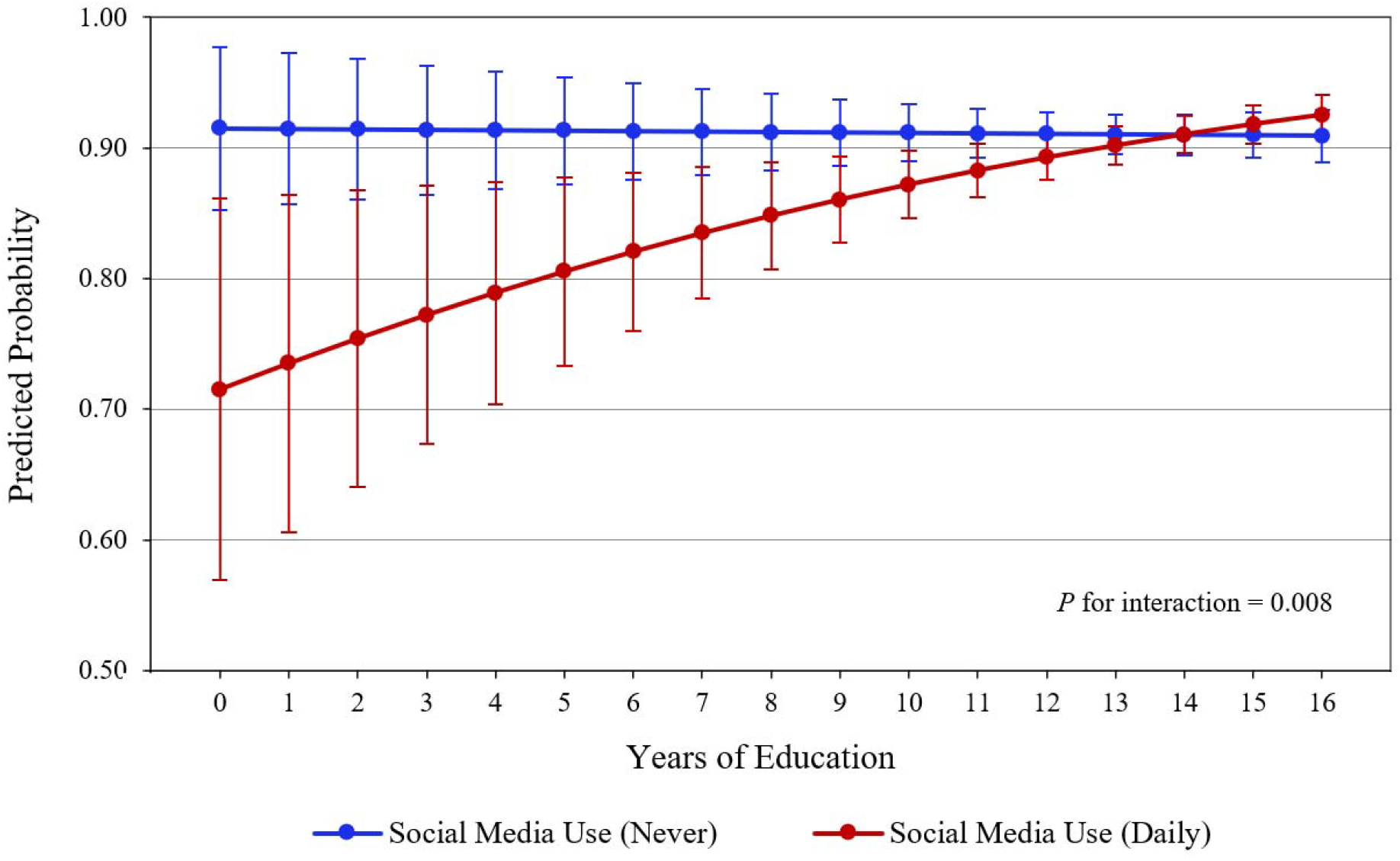
Predicted Probability of Covid-19 Vaccination by Social Media Use and Education, Health and Retirement Study, 2022 (n=4,038) *Note*: Estimates obtained from Table 2 (Model 3) adjusting for age, sex, race, ethnicity, geographic region, marital status, religious attendance, frequency of computer use, frequency of internet use for health information, and reported diagnoses of high blood pressure, diabetes, heart disease, and cancer.

## DISCUSSION

Our findings from a national cohort showed a complex association between social media use and vaccination for Covid-19 among U.S. middle-aged and older adults. Results showed that social media use was not directly associated with getting vaccinated for Covid-19. However, we found that the association between social media use and vaccination varied significantly by years of educational attainment. In adults who frequently used social media, results showed that those with low education were significantly less likely to get vaccinated for Covid-19. Furthermore, we found that social media use among middle-aged and older adults did not influence being vaccinated for influenza, pneumonia, or shingles.

The central finding that less-educated frequent-users of social media were less likely to receive Covid-19 vaccination suggests that social media may be a conduit for vaccine hesitancy among vulnerable populations with lower educational attainment. This finding aligns with previous research documenting the spread of Covid-19 vaccine misinformation on major social media platforms.^10^ This also suggests that adults with lower levels of education—and likewise health literacy—may be more susceptible to such content.^9^ Furthermore, the specificity of the association to Covid-19 vaccination, and not with other vaccinations (i.e., influenza, pneumonia, etc.), likely reflects the widespread politicization and disinformation surrounding vaccination for Covid-19 in the United States. Unlike more established vaccines familiar to middle-aged and older adults, vaccination for Covid-19 became a lightning-rod for intense debate and the spread of misinformation campaigns.^15^ Furthermore, the “shelter-in-place” policies that were enacted, and the increasing culture of remote work that followed, likely amplified the role of social media in the (mis)communication of health information during this time.

This study had several limitations that should be acknowledged. First, we could not directly assess the specific content and/or quality of health information encountered on social media. Similarly, social media use was measured broadly across platforms (Facebook, Instagram, etc.) and did not distinguish between passive consumption and active engagement with the content. The limited sample size also precluded additional analyses of potentially meaningful differences in population subgroups (e.g., among demographic subgroups, persons with pre-existing health conditions, etc.). Finally, the analyses were based on cross-sectional data that prevented any causal interpretations.

In summary, the results of this report highlight the importance of delivering tailored public health messages when promoting vaccine uptake among less educated and vulnerable older populations. Public health agencies and health care providers should consider individuals’ educational attainment when designing and deploying communication strategies to counter the potential misinformation on social media sites that can be widely consumed by middle-aged and older adults.

## Data Availability

All data produced in the present study are available upon reasonable request to the authors

