## Supplemental Tables for "Digital Divide or Educational Divide? The Impact of Social Media on COVID-19 Vaccination in Middle-Aged and Older Adults"

| **Supplementary Table 1.** Model Estimates of the Association Between Social Media Use, Education, and Vaccination for Influenza, HRS 2022 (n=4,038) | | | | | | |
| --- | --- | --- | --- | --- | --- | --- |
|  | **Model 1** | | **Model 2** | | **Model 3** | |
|  | OR [95% CI] | *P*  value | OR [95% CI] | *P*  value | OR [95% CI] | *P*  value |
| Frequency of Social Media Use | 0.98 [0.94-1.02] | .336 | 1.02 [0.97-1.07] | .397 | 0.98 [0.80-1.18] | .806 |
| Years of Education |  |  | 1.03 [1.00-1.06] | .076 | 1.02 [0.98-1.06] | .341 |
| Interaction Term |  |  |  |  |  |  |
| Social Media Use * Education |  |  |  |  | 1.00 [0.99-1.02] | .638 |
| Abbreviations: HRS, Health and Retirement Study; OR, odds ration; CI, confidence interval.  *Note*: Models 2 and 3 adjusted for age, sex, race, ethnicity, geographic region, marital status, religious attendance, frequency of computer use, frequency of internet use for health information, and reported diagnoses of high blood pressure, diabetes, heart disease, and cancer. | | | | | | |

| **Supplementary Table 2.** Model Estimates of the Association Between Social Media Use, Education, and Vaccination for Shingles, HRS 2022 (n=4,038) | | | | | | |
| --- | --- | --- | --- | --- | --- | --- |
|  | **Model 1** | | **Model 2** | | **Model 3** | |
|  | OR [95% CI] | *P*  value | OR [95% CI] | *P*  value | OR [95% CI] | *P*  value |
| Frequency of Social Media Use | 1.05 [1.00-1.10] | .071 | 0.99 [0.94-1.05] | .752 | 0.98 [0.78-1.24] | .867 |
| Years of Education |  |  | 1.02 [0.99-1.05] | .263 | 1.02 [0.97-1.07] | .480 |
| Interaction Term |  |  |  |  |  |  |
| Social Media Use * Education |  |  |  |  | 1.00 [1.98-1.02] | .925 |
| Abbreviations: HRS, Health and Retirement Study; OR, odds ration; CI, confidence interval.  *Note*: Models 2 and 3 adjusted for age, sex, race, ethnicity, geographic region, marital status, religious attendance, frequency of computer use, frequency of internet use for health information, and reported diagnoses of high blood pressure, diabetes, heart disease, and cancer. | | | | | | |

| **Supplementary Table 3.** Model Estimates of the Association Between Social Media Use, Education, and Vaccination for Pneumonia, HRS 2022 (n=4,038) | | | | | | |
| --- | --- | --- | --- | --- | --- | --- |
|  | **Model 1** | | **Model 2** | | **Model 3** | |
|  | OR [95% CI] | *P*  value | OR [95% CI] | *P*  value | OR [95% CI] | *P*  value |
| Frequency of Social Media Use | 1.06 [0.96-1.16] | .246 | 1.05 [0.94-1.18] | .560 | 1.08 [0.52-2.22] | .826 |
| Years of Education |  |  | 1.04 [0.98-1.11] | .186 | 0.99 [0.86-1.14] | .928 |
| Interaction Term |  |  |  |  |  |  |
| Social Media Use * Education |  |  |  |  | 0.99 [0.94-1.05] | .803 |
| Abbreviations: HRS, Health and Retirement Study; OR, odds ration; CI, confidence interval.  *Note*: Models 2 and 3 adjusted for age, sex, race, ethnicity, geographic region, marital status, religious attendance, frequency of computer use, frequency of internet use for health information, and reported diagnoses of high blood pressure, diabetes, heart disease, and cancer. | | | | | | |
